# Electronic Nicotine Delivery Systems Use and Reduced Days of Cigarette Smoking: A Causal Bayesian Additive Regression Tree Analysis of Population Assessment of Tobacco and Health Study Waves 4 – 6

**DOI:** 10.64898/2026.11.23.26368803

**Authors:** Shu Xu, Jennifer Hill, Luchang Cui, Yang Feng, Raymond S. Niaura

## Abstract

**Background:** Debate persists regarding the potential harm reduction role of Electronic Nicotine Delivery Systems (ENDS). This study estimated the effect of ENDS use on subsequent cigarette smoking days in the past 30 days among adult smokers.

**Methods:** We analyzed longitudinal data from Waves 4 to 6 of the Population Assessment of Tobacco and Health (PATH) Study, including 4,909 established current smokers. Bayesian Additive Regression Trees for causal inference was used to estimate the average exposure effect (ATE) of current ENDS use (characterized as no use, some-day use, and daily use) at Wave 5 (2018 - 2019) on cigarette smoking days at Wave 6 (2021), adjusting for 26 covariates measured at Wave 4 (2016 - 2018).

**Results:** Compared with no ENDS use, daily ENDS use reduced cigarette smoking by 6.17 days per month over a two-year period (ATE = −6.17, 95% CI: −8.16 to −5.29); whereas some-day use reduced smoking by 0.99 days (ATE = −0.99, 95% CI: −1.65 to −0.28). Compared with some-day use, daily ENDS users reduced smoking by 5.65 days per month (ATE = −5.65, 95% CI: −6.72 to −4.72). There was no evidence that these effects differed meaningfully across demographic or psychosocial subgroups.

**Conclusions:** Among established adult smokers, daily ENDS use reduced subsequent smoking days compared with some-day and no ENDS use. These findings support the potential role of ENDS as a harm reduction strategy for adult smokers while highlighting the need for better characterizing behavioral patterns and health consequences associated with sustained ENDS use.

**Implications:** Among US adult established current smokers, Daily ENDS use reduced cigarette smoking by approximately 6 days per month over a two-year period compared with some-day and no ENDS use. These effects do not vary meaningfully by sex, education, race/ethnicity, annual household income, mental health status, sexual orientation, or age, suggesting consistent impacts across diverse populations. Findings support the potential of ENDS as a harm reduction tool for adult smokers and warrents more understanding on sustained dual smokers who vaps.

- BART effectively addresses high-dimensional and potentially interactive confounding.

## Effects of Electronic Nicotine Delivery Systems Use on Reduced Cigarette Smoking Days

Cigarette smoking remains prevalent in the United States (US) and is the leading cause of preventable morbidity and mortality.^1^ Smoking cessation therefore remains a public health priority.^2^ Although evidence-based cessation interventions and U.S. Food and Drug Administration (FDA)-approved pharmacotherapies are available, their uptake is limited and quit rates remain modest (e.g., 8.8% in 2022),^3–5^ underscoring the need for alternative accessible and effective strategies.

Electronic Nicotine Delivery Systems (ENDS), commonly known as e-cigarettes, have emerged as a potential harm-reduction and cessation tool.^9^ ENDS deliver nicotine via aerosol without the products of tobacco combustion and are therefore generally considered less harmful than cigarettes.^6–10^ ENDS have become increasingly accessible and widely used among U.S. adults, with current use rising from 1.9% in 2012 to 7.0% in 2024, despite a temporary decline to 3.7% in 2020.^11–15^ Most adult ENDS users are current or former cigarette smokers, particularly younger adults,^16^ and many report using ENDS to reduce or quit smoking. Randomized clinical trials have provided growing evidence supporting the efficacy of ENDS for smoking cessation,^9^ whereas findings from observational studies remain mixed.^17^ This discrepancy suggests that further research is needed to assess the efficacy of ENDS on cigarette smoking under real-world conditions, across multiple smoking-related outcomes, and both in the overall population and socio-demographically diverse subpopulations. Notably, a recent review of 38 analyses found that ENDS use – particularly daily use compared with some-day or no use – was associated with a greater likelihood of smoking cessation. ^17^

Assessing the efficacy of ENDS use on cigarette smoking using observational data is challenging because ENDS exposure is not randomly assigned. Thus, exposed and unexposed individuals may differ in ways that influence the outcomes.^18, 19^ To combat this potential bias, previous studies have adjusted for a variety of potential confounders, including sociodemographic, interpersonal, behavioral, and social environmental factors, though the specific confounders included have varied considerably across studies. Recent developments in modern causal inference methods, including the Bayesian Additive Regression Trees (BART) approach, provide a principled framework for estimating causal effects from observational data by explicitly accounting for observed confounding under a set of well-defined assumptions.^20,21^

Another challenge in assessing the efficacy of ENDS is the choice of outcome. Smoking behavior is commonly quantified using smoking frequency, the number of days smoked in the past 30 days (range: 0 to 30), hereafter referred to as smoking days. However, in the scientific literature, when used as an outcome variable, it is often dichotomized (e.g., splitting at a cut-point such as none vs any use in the past 30 days) or categorized into small groups.^17^ Key limitations of this approach include several well-documented statistical drawbacks^22,23^: loss of information, as collapsing a continuous measure removes fine-grained variation and reduces analytic sensitivity and efficiency; reduced statistical power, increasing risk of false-negative findings; and potential distortion of effect estimates, as evidenced in simulation studies.

Although smoking days is inherently a continuous variable, it has rarely been analyzed as such. This outcome often exhibits clustering at both the lower and upper bounds, reflecting participants who quit smoking (0 days) and those who continue to smoke daily (30 days). The bounded and bimodal nature of this distribution complicates standard continuous-outcome modeling.^24^

To our knowledge, our study is among the first to estimate the efficacy of ENDS use on subsequent smoking days. Using longitudinal observational data from Waves 4 – 6 (2016 – 2021) of the Population Assessment of Tobacco and Health (PATH) Study, we analyzed a cohort of established adult smokers and applied BART for causal inference to account for potential confounding by factors associated with both ENDS use and smoking behaviors.^20^ Specifically, we addressed the following research questions.

### Q1 (Average Exposure/Treatment Effect; ATE)

What would be the expected change in the average number of subsequent smoking days over a two-year follow-up period if smokers had used ENDS daily (i.e., daily ENDS use vs. no use) or on some days (i.e., some-day ENDS use vs. no use) compared to no ENDS use?

### Q2 (Group-specific Average Exposure/Treatment Effect; Group-specific ATE)

Are any effects identified for Q1 moderated by participants’ sex, education, race/ethnicity, annual household income, internalization, externalization, sexual orientation, or age?

## Methods

### Data source and study sample

The PATH Study is a nationally representative longitudinal cohort supported by the National Institutes of Health (NIH) and the FDA to inform tobacco regulatory science by characterizing population-level tobacco use patterns and associated health outcomes following the expansion of FDA regulatory authority in 2009. Since 2013, PATH has followed youth (ages 12–17 years; n = 13,651) and adults (ages ≥18 years; n = 32,320), including tobacco users and non-users, across multiple waves. Each wave collects extensive information on tobacco and nicotine product use, dependence, transitions in use, and health indicators, providing a comprehensive resource for studying tobacco behaviors and their context factors (see published design documentation for details).^25^

In this study, we included 4,909 established current cigarette smokers who met the following criteria: a) participated in Waves 4 (12/2016-01/2018; Baseline wave), 5 (12/2018 – 11/2019; Exposure wave), and 6 (03/2021 – 11/2021; Outcome wave) data collection; therefore they have non-missing values on sampling weights for primary analyses; b) reported having smoked more than 100 cigarettes in their lifetime at baseline and had smoked within the past 30 days at both the baseline and exposure waves; c) reported ENDS use status at the exposure wave, d) no missing values on all baseline covariates; and e) reported their cigarette smoking days at the outcome wave. The participant inclusion flowchart is presented in Figure S1.

### Measures

To facilitate causal interpretation, we enforced a strict temporal ordering of variables, such that covariates precede exposure and exposure precedes the outcome. This ordering ensures that causes temporally precede effects and mitigates issues arising from adjusting for potential mediators, colliders, or bidirectional relationships, which can bias the causal effect estimate.^26,27^ Details on study variables are presented in Table S1.

**ENDS use exposure** (Wave 5) was measured using two self-reported items. Participants were first asked whether they had used ENDS in the past 30 days. Those who reported “Yes” were then asked whether they had used ENDS not at all, on some days, or every day during that period. Based on these responses, ENDS use was categorized as no (current) use [reference group], some-day use, and daily use.

**Smoking days outcome** (Wave 6). Similar to ENDS use, participants were asked whether they had smoked cigarettes in the past 30 days. Those who reported “Yes” were subsequently asked whether they had smoked not at all, on some days, or every day during that period. Only participants who reported some-day smoking were asked to indicate the number of smoking days in the past 30 days. Those who reported daily smoking or not at all were not asked the follow-up question about the number of smoking days; therefore, we assigned a value of 30 days to indicate daily smoking and a value of 0 days to indicate no smoking. Therefore, smoking days is a numerical value ranging from 0 to 30.

**Covariates** (Wave 4). We adjusted for 26 pre-exposure covariates (22 categorical and 4 continuous variables), as they are potentially associated with both ENDS use exposure and subsequent smoking days.^17,28,29^ Pre-exposure covariates included age group, sex, race/ethnicity, educational attainment, annual household income, region, sexual orientation, absolute harm perception of cigarette smoking, relative harm perception of ENDS use compared to cigarette smoking, house policy of cigarette use, family tobacco use, internalization, externalization, heart disease, respiratory disease, or cancer in the past 12 months, body mass index; past month marijuana use and alcohol use; other combustible tobacco use, cigarette quit intention, secondhand cigarette smoking exposure in the past 7 days, time to first smoking after waking, and nicotine dependence. Covariates also included baseline measures of exposure and outcome variables. Missing values for categorical covariates were recoded as a “Don’t know” category. For the BART analyses, categorical variables were converted into dummy variables, resulting in a total of 64 dummy variables and four continuous covariates.

### Analytic Strategy

Descriptive statistics of study variables were reported for the overall sample and by ENDS exposure level. To examine the effect of ENDS use on subsequent smoking days over a two-year period, we used BART for causal inference for the primary analyses and performed sensitivity tests to evaluate the impact of potential violations of key assumptions. Based on prior studies,^17^ our primary research questions focus on two comparisons: Daily ENDS use versus No use (n = 4303), and Some-day ENDS use versus No use (n = 4638). For completeness, we also estimated the effects of Daily ENDS use versus Some-day use (n = 877). Accordingly, we conducted three parallel analyses, with each exposure comparison estimated in separate analyses.

BART is a flexible, nonparametric modeling approach that integrates boosted regression tree models within a Bayesian inferential framework.^30^ It enables flexible modeling of an outcome variable conditional on a set of covariates, without imposing parametric assumptions. It constructs an ensemble of many small regression trees, where each tree makes a small contribution to the overall fit. This structure helps capture complex, nonlinear relationships between the outcome and the covariates, while effectively mitigating overfitting through three weakly informative priors. BART is fit using Markov Chain Monte Carlo methods.

### BART for causal inference

BART has gained popularity for both its statistical development and its application to causal inference research.^20,21,31–33^ A key advantage of BART over alternative methods (e.g., propensity score approaches) for causal inference is the fact that it doesn’t require the iterative process of model fitting, data restructuring (for instance matching or weighting), and balance checking across multiple cycles that has the disadvantage of complicating the procedure and increasing “researcher degrees of freedom.”^34,35^ BART has demonstrated superior performance in numerous complicated inferential settings including several causal inference data analysis challenges. ^33,36,37^

BART for causal inference methods is more easily conceptualized within the potential outcomes framework,^38^ which posits two potential outcomes in response to a binary exposure: *Y*(1), the outcome that would be observed under exposure, and *Y*(0), the outcome that would be observed absent the exposure. Because only one of these potential outcomes can be observed for any participant, the other is counterfactual (or, potential). BART estimates causal effects by flexibly modeling the response surface, that is the relationship between pre-exposure covariates, exposure assignment, and the outcome, and using this model to predict counterfactual outcomes. Technical details of the BART for causal inference model are provided in the Appendix and described elsewhere.^20,21,39,40^

#### Estimands

In our study, BART was used to make predictions (along with associated uncertainty) for a participant’s potential outcomes under three distinct scenarios: daily ENDS use, some-day ENDS use, or no ENDS use in the past 30 days, respectively. The participant’s causal effect for the comparison of any two of these exposure regimens was defined as the expected difference between the two corresponding potential outcomes. The estimands corresponding to Q1 take the average of these differences over all participants, whereas those for Q2 focus on specified subgroups.

#### Identification assumptions

A causal interpretation of the estimated exposure effect relies on several key assumptions necessary for the identification of these causal estimands.^20^ First, we assumed *no unmeasured confounders* (also referred to as *ignorability* or *selection on observables*). Given the extensive set of covariates included in the analysis, we believe that this assumption is plausible, ensuring that potential outcomes are independent of exposure assignment when conditioned on covariates. Second, we assumed the *positivity* condition (also referred to as *overlap* or *common support*), which requires that all individuals have a nonzero probability of receiving each exposure level across all levels of the covariates, allowing for meaningful causal comparisons. Finally, we assumed the *Stable Unit Treatment Value Assumption* (consisting of *consistency* and *no interference*), meaning that a participant’s outcome depends solely on one’s own exposure status, not influenced by the exposure status of others. It also requires that everyone receives the same version of the exposure. The assumption is more plausible if we consider broad definitions of the exposure (for instance, inclusive of different versions of ENDS and ignoring distinctions in amounts of consumption within exposure categories).

Unlike inverse propensity score weighting or propensity score matching, BART adjusts for confounding through flexible outcome modeling rather than by restructuring datasets to try to create comparability across groups. Therefore, covariate balance diagnostics commonly used to evaluate the comparability of samples in those approaches are not required here. However, automated common-support diagnostics were implemented to assess positivity.

#### Estimation

In this study, we fit BART models with a continuous outcome under the potential outcomes framework for each of two research questions. Each model predicts potential outcomes conditional on both observed covariates and an estimate of the propensity score. Including a propensity score estimate in the model for the exposure effect has been shown to yield more reliable results as a way to control for confounding.^41^ For each research question, BART generated a posterior distribution of individual-level potential outcomes at each exposure level conditional on the covariates of these participants. For each analysis, we ran 10 independent Markov Chains with 200 burn-in iterations and 500 sampling iterations per chain, fixing the number of trees in the ensemble to 200. Trace plots were examined to ensure convergence of chains. A point estimate of ATE was obtained as posterior mean across retained MCMC posterior draws over the whole sample (Q1) and each specific subgroup (Q2). The uncertainty of these estimates were obtained through analyses using the resampling weights (see the Weighted Analysis section).

Because the outcome is continuous, potential outcomes were expressed in (smoking) days, with negative ATE estimate indicating reduced smoking days.

#### Diagnostics

To assess the adequacy of BART models with respect to the challenging outcome distribution, we examined the distribution of the residuals. Residual distributions that were approximately symmetric and exhibited an absolute skewness value below 3 were considered indicative of an adequate model fit.

#### Sensitivity analysis

To assess the robustness of ATE estimates to potential unmeasured confounding, we assumed the existence of a hypothetical unobserved confounder and characterized its associations with the exposure and outcome using two sensitivity parameters.^42,43^ ATEs were subsequently re-estimated across a range of plausible combinations of treatment–confounder and outcome–confounder associations. Results were summarized using contour plots, which display the estimated exposure effect under varying assumptions regarding the strength of unmeasured confounding.

### Weighted analysis

Sampling weights (all-waves weights: *R06_A_S04WGT*) were incorporated into primary analyses for the estimates of causal effects. To quantify uncertainty, we repeated the estimation procedure 100 times using the PATH replicate weights and constructed 95% credible intervals (CI) from the empirical distribution of the resulting estimates, following guidance from the PATH Technical Working Group. Sampling weights accounted for the multiple-stage sampling design, adjusted for attrition, and supported valid population inference.^44^ Weighted sensitivity tests are not feasible and will require advances in methodology and software packages.

### Missing values

For categorical covariates, missing values were treated as a separate (“Don’t know”) category. Participants with missing values on any other study variables were excluded using complete-case analysis (3.8%), as the proportion of missing data was small and assumed to be ignorable.

### Software

BART analysis for causal inference was conducted using the R *bartCause* package. Sensitivity analyses were conducted using the R *treatSens* package.^42,43^

## Results

Descriptive statistics for baseline characteristics are presented for the overall study sample and stratified by ENDS exposure status in Table S2. Participants were mainly between 18 and 64 years old (95.6%), with approximately half female (54.2%) and about three-fifth as non-Hispanic White (62.8%). All participants were established current cigarette smokers at both the baseline and exposure waves.

At the exposure wave, 5.5% (n = 271) of participants reported daily ENDS use, 12.3% (n = 606) reported some-day ENDS use, and 82.1% (n = 4032) reported no (current) ENDS use. At the outcome wave, participants who had reported daily ENDS use exhibited the highest prevalence of zero smoking days (35.4%, n = 96) and the lowest prevalence of daily cigarette smoking (43.5%, n = 118), followed by some-day ENDS users (16.0%, n = 97; 67.3%, n = 408, respectively) and non-users (12.7%, n = 512; 75.6%, n = 3050, respectively). Consistent with these patterns, daily ENDS users had the lowest mean and median number of smoking days (mean = 14.87, SD = 14.09; median = 10), followed by some-day ENDS users (mean = 22.07, SD = 12.27; median = 30), whereas non-users had the highest values (mean = 24.02, SD = 11.24; median = 30). Figure 1 presents a histogram of the outcome variable.

**Figure 1.**
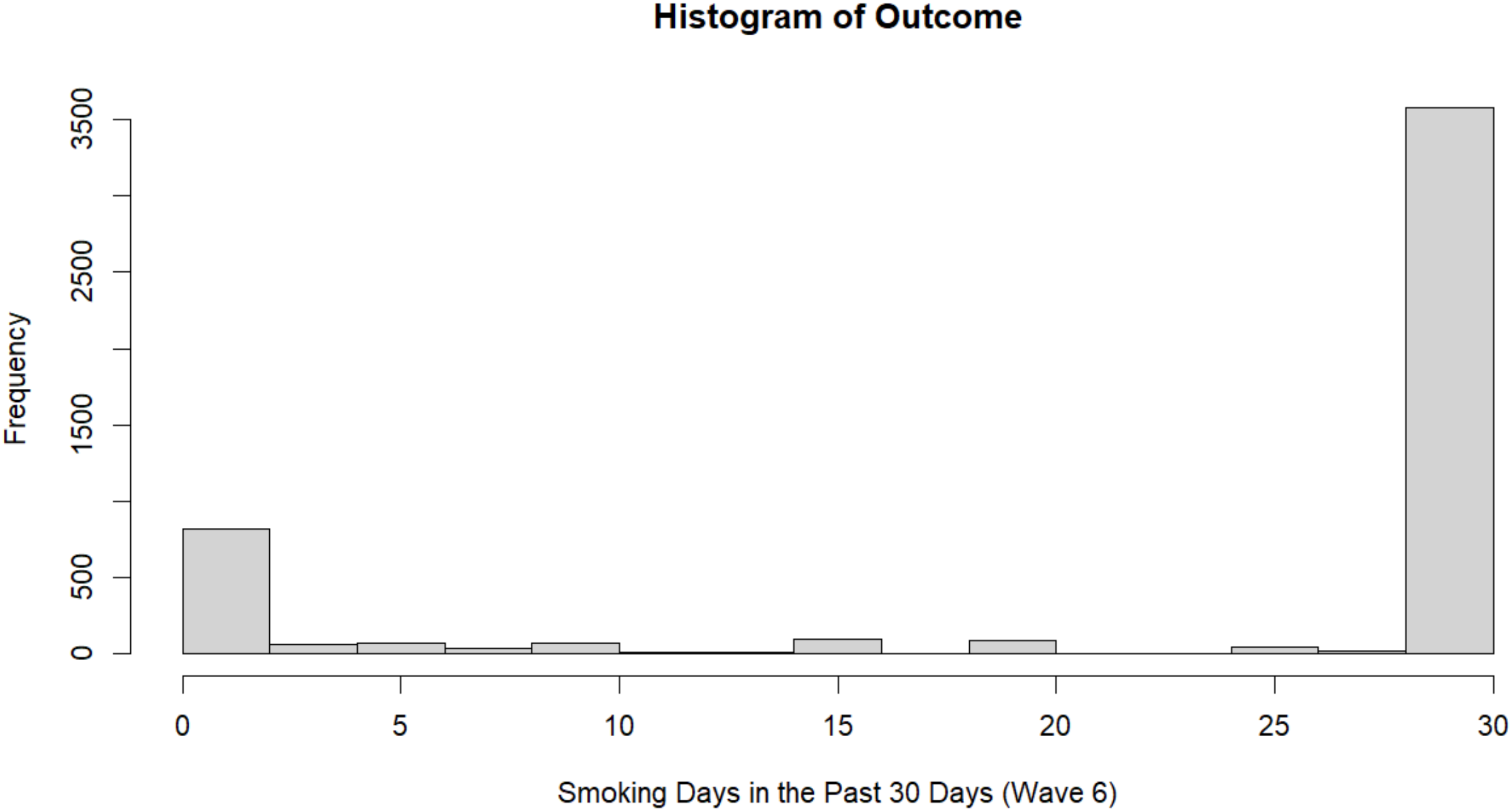
Histogram of the Cigarette Smoking Days at the Outcome Wave

### Q1: ATE

As shown in Figure 2 and Table S3, compared with no ENDS use, daily ENDS use reduced cigarette smoking by 6.17 days per month over a two-year period (ATE = −6.17, 95% CI: −8.16 to −5.29); whereas some-day use reduced smoking by 0.99 days (ATE = −0.99, 95% CI: −1.65 to −0.28). Compared with some-day use, daily ENDS use reduced smoking by 5.65 days per month (ATE = −5.65, 95% CI: −6.72 to −4.72). Across all analyses, only 0 to 2 participants were excluded to satisfy the common support requirement. In addition, residuals were approximately normally distributed, with absolute skewness values below the prespecified threshold (Table S4), indicating that the BART models provided an adequate fit to the data.

**Figure 2.**
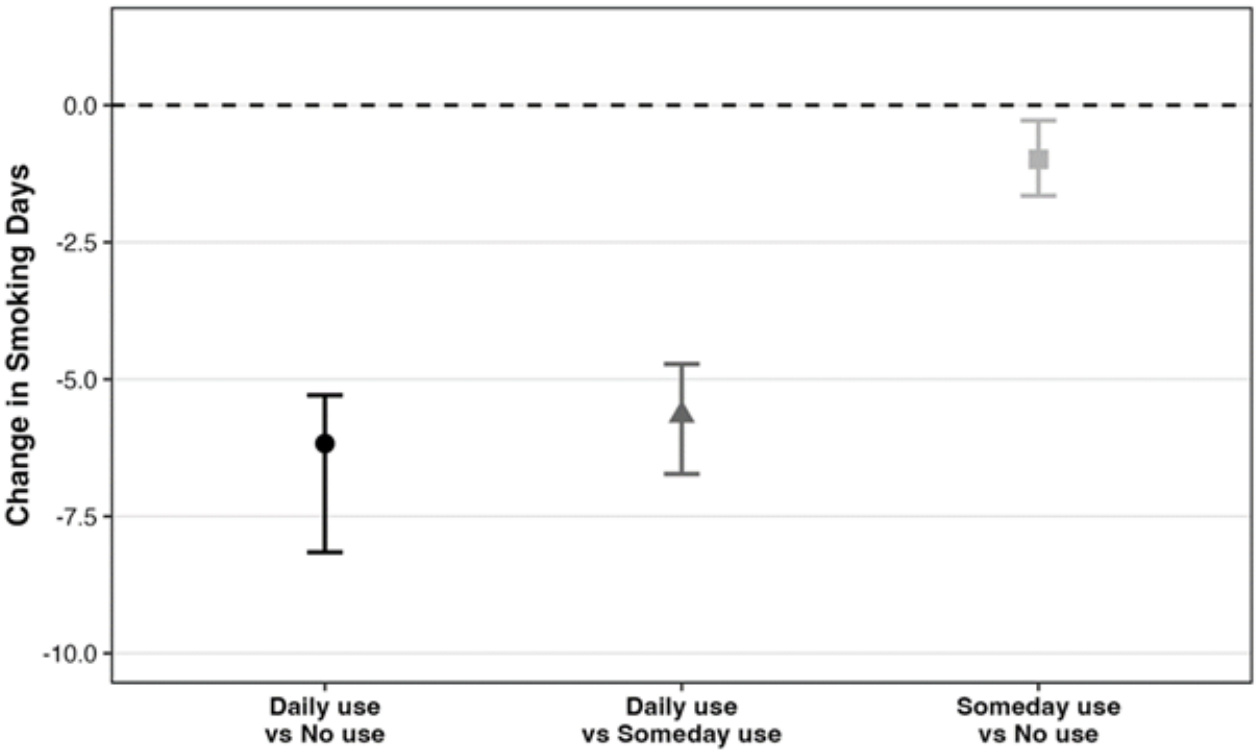
Estimate of Average Exposure Effect of ENDS Use on Subsequent Smoking Days in the Overall Population

### Q2: Group-Specific ATE

We further examined subgroups and found no strong evidence that the group-specific ATE of ENDS use varied meaningfully by sex, race/ethnicity, annual household income, externalization, internalization, sexual orientation, or age group. As shown in Figure 3 and Table S3, subgroup-specific estimates exhibited substantial overlap in their 95% credible intervals, suggesting generally consistent ENDS exposure effects across subgroups. The only potential exception was observed in older age groups with extremely small sample size, where the estimates were less precise and less robust.

**Figure 3.**
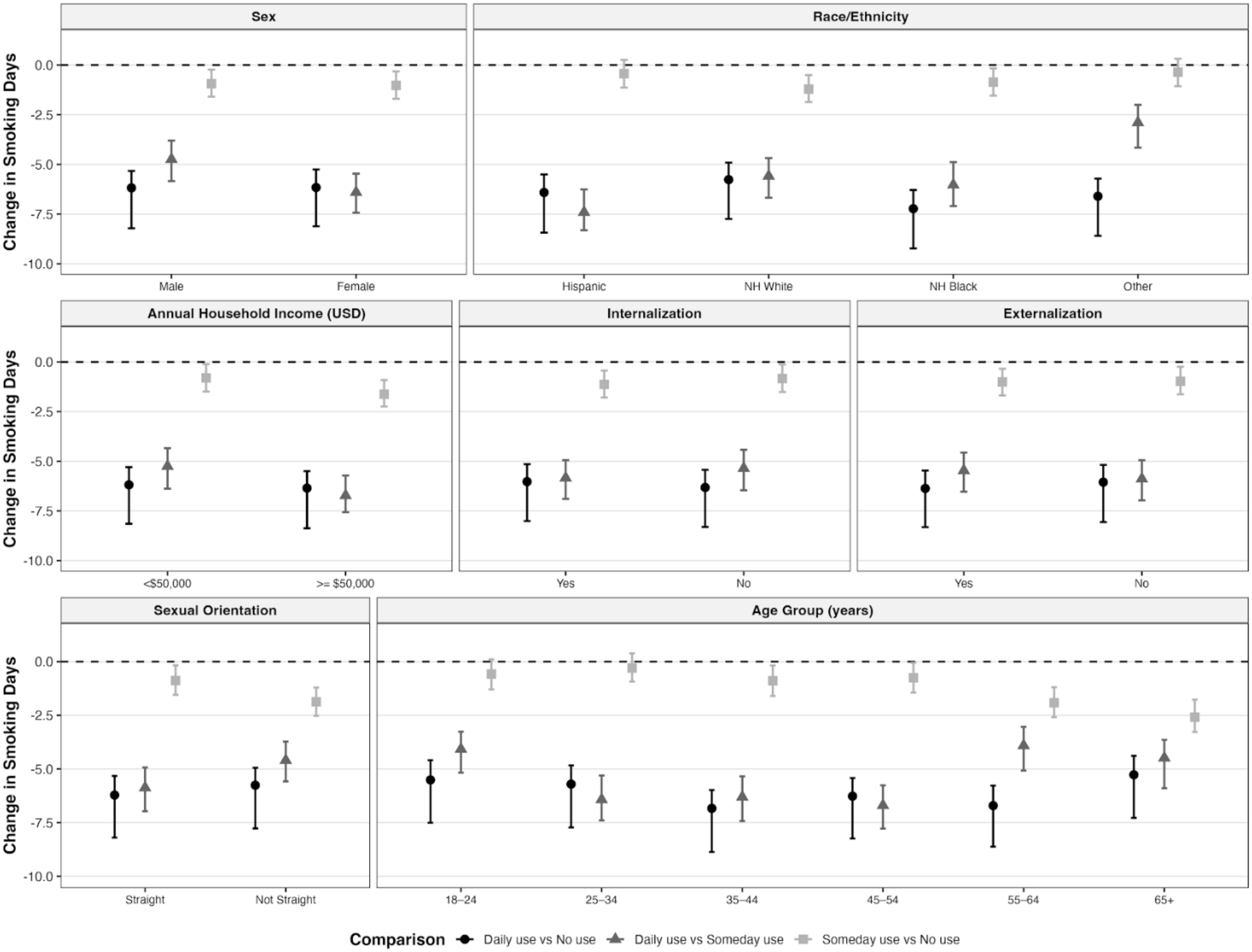
Estimate of Group-Specific Average Exposure Effect of ENDS Use on Subsequent Smoking Days Within Population Subgroups.

## Sensitivity analysis

Sensitivity analyses suggested that the estimated ATEs were relatively insensitive to moderate levels of unmeasured confounding. Across a wide range of plausible exposure-confounder and outcome-confounder associations, the ATEs remained negative and of similar magnitude. Furthermore, the observed confounders clustered in regions where the estimates changed minimally, suggesting that an unmeasured confounder would need to be substantially stronger than most measured confounders to meaningfully alter the study conclusions (Figure S2).

## Discussion

To the best of our knowledge, this is among the first studies to apply a causal machine learning approach, BART for causal inference, to estimate the causal effect of ENDS use on subsequent smoking days while accounting for high-dimensional confounding. Using longitudinal data from the nationally representative PATH Study, we found that ENDS use reduced smoking days over a two-year follow-up among established adult smokers. Compared with non-users, daily ENDS use reduced smoking by approximately 6 days per month, whereas some-day ENDS use reduced smoking by about 1 day per month. These findings are consistent with previous studies reporting that daily ENDS use was associated with cigarette cessation,^17^ while extending the literature by quantifying reductions in smoking days. Moreover, these findings were consistent among younger age, race/ethnicity, sex, household income, education, mental health status, or sexual orientation subgroups.

Unlike traditional regression approaches, which primarily estimate associations between exposures and outcomes, our study was conducted within the potential outcomes framework,^38,45^ in which causal effects are defined as contrasts between potential outcomes under alternative exposure conditions. For example, we estimated the expected difference in smoking days had smokers used ENDS daily rather than on some days. This framework provides a rigorous and transparent way of defining, estimating, and interpreting causal effects while explicitly distinguishing causal questions from associational ones.^39^ Identification of these causal effects relied on several assumptions, including no unmeasured pre-exposure confounding, positivity, and no interference between participants. To strengthen the plausibility of these assumptions, we adjusted for a comprehensive set of baseline covariates. Sensitivity analyses further indicated that an unmeasured confounder would need to be substantially stronger than the measured confounders to meaningfully explain away the effect of daily ENDS use. Finally, given the PATH Study design and the geographic dispersion of participants, interference between participants was considered unlikely.

BART provides a flexible means for estimating causal effects from observational data. Unlike traditional regression models, BART accommodates complex nonlinear relationships and high-order interactions without requiring investigators to prespecify interaction terms, transformations, or functional forms, thereby reducing the risk of model misspecification and limiting the researcher degrees of freedom. In the *bartCause* package, causal effect estimation is further strengthened by incorporating estimated propensity scores. Unlike many machine learning algorithms designed primarily for prediction,^46^ BART is implemented within a Bayesian framework, allowing uncertainty in causal effect estimates to be quantified through posterior distributions and credible intervals. BART also combines an ensemble of many small regression trees with regularizing priors, which reduces overfitting while preserving the flexibility needed to model complex exposure–outcome relationships. Together, these features enable robust estimation of both population-average and subgroup-specific causal effects while minimizing reliance on restrictive modeling assumptions.

While the bimodal distribution of smoking days presents potential presents modelling challenges, the BART models appear to provide an adequate fit to the data. This approach allowed us to analyze smoking days as a continuous outcome, rather than characterizing smoking behavior into two (any vs. no smoking) or ordinal (daily vs. some-day vs. no smoking) categories.

Our findings add to the relatively limited literature examining smoking days as an outcome and have important implications for the potential harm reduction role of ENDS. Although complete abstinence from all tobacco and nicotine products remains the ideal public health goal,^2^ reducing the use of combustible cigarettes may represent an intermediate step toward lowering tobacco-related harm. In this study, daily ENDS use reduced smoking days by approximately 6 days per month over a two-year period, suggesting that some smokers may substitute ENDS for combustible cigarettes. Nevertheless, these findings should be interpreted with caution. Fewer smoking days do not necessarily translate into proportional reductions in health risks, particularly among individuals who continue smoking cigarettes or engage in dual use of cigarettes and ENDS.^47^ The long-term health consequences of sustained dual use remain uncertain and may include continued exposure to cigarette smoke,^47^ persistent nicotine dependence, delayed tobacco abstinence, and cumulative adverse health effects.^48^ Accordingly, our findings should not be interpreted as evidence that ENDS are harmless or that switching to ENDS eliminates health risks associated with tobacco use. Importantly, individuals who do not currently use tobacco products, especially youth and tobacco-naïve adults, should not initiate ENDS or any other tobacco product.

This study has several limitations. First, causal interpretation depends on the assumption of no unmeasured confounders. Although we adjusted for a comprehensive list of pre-exposure covariates selected based on the literature and subject-matter knowledge, it is possible that some critical factors were omitted. Second, ENDS exposure was assessed at a single time point and categorized by frequency of use; we were unable to evaluate the effects of nicotine dose, product characteristics, or cumulative ENDS use over time. Third, the study conclusions should only be generalized with caution. At the time of exposure assessment (12/2018 – 11/2019), the US ENDS market was dominated by pod-based ENDS products – particularly JUUL--which marked a significant shift in vaping technology and use patterns.^49^ These devices differ substantially from the earlier devices (e.g., cigalike and box mods with slower and lower nicotine delivery) and newer products (2020s – present), which integrate smart technology such as Bluetooth connectivity, touchscreen interfaces, and app compatibility for personalized settings.^50^ The effect of ENDS use on smoking cessation likely varies by device design, nicotine delivery, and therefore, user behavioral patterns. Further direct comparative research is warranted. Our findings should not be generalized beyond our target population, study period, and geographic setting.

## Conclusion

Among established adult smokers, daily ENDS use reduced subsequent smoking days compared with some-day or no use. These findings support the potential role of daily ENDS use in tobacco harm reduction while underscoring the need to evaluate its long-term health effects and identify the smokers most likely to benefit.

## Data Availability

This study used the Public Use data from the Population Assessment of Tobacco and Health Study. The data (ICPSR 36498) are publicly available at the Inter-university Consortium for Political and Social Research. https://doi.org/10.3886/ICPSR36498.v19.

## Supporting information

Supplementary All

## Data Availability

The data (ICPSR 36498) are publicly available at the Inter-university Consortium for Political and Social Research. https://doi.org/10.3886/ICPSR36498.v19.

