## Supplementary All for "Electronic Nicotine Delivery Systems Use and Reduced Days of Cigarette Smoking: A Causal Bayesian Additive Regression Tree Analysis of Population Assessment of Tobacco and Health Study Waves 4 – 6"

Supplementary Materials

Table S1 Description of Study Variables……………………………………………………………………………………….. 2
Table S2 Characteristics of the study sample at Wave 4 and by Exposure Groups……………………………………….……….13
Table S3 Causal Effects ENDS Use on Subsequent Smoking Days: Estimates of ATE and Group-Specific ATE……...……………..11
Table S4 Residual Distribution Statistics from BART for ATE analyses. ...…...………………………………………………………14

Figure S1 Flowchart of Sample Selection…………………………………………………………………………………...…..16
Figure S2 Contour Plots of Sensitivity to unmeasured confounding for ATEs……………………………………..…………………17

**Appendix**. An introduction to Bayesian Additive Regression Tree for Causal Inference: a Causal Machine Learning Method

Table S1
Description of Study Variables

| | PATH Variable | Label | Response Category | Recoded Response Category |
|---|---|---|---|---|
| | | **Covariates (Wave 4)** | | |
| Age Group | R04R_A_AGECAT6 | Age (in years) at the time of interview. | 1 = 18 to 24 years old;<br>2 = 25 to 34 years old;<br>3 = 35 to 44 years old;<br>4 = 45 to 54 years old;<br>5 = 55 to 64 years old;<br>6 = 65 years old or older | 1 = 18 to 24 years old;<br>2 = 25 to 34 years old;<br>3 = 35 to 44 years old;<br>4 = 45 to 54 years old;<br>5 = 55 to 64 years old;<br>6 = 65 years old or older |
| Sex | R04R_A_SEX_IMP | PATH imputed gender | 1 = Male;<br>2 = Female | 0 = Female;<br>1 = Male |
| Race/Ethnicity | R04R_A_RACECAT3 | Race | 1 = White alone;<br>2 = Black alone;<br>3 = Other | (recoded into four binary variables)<br>Hispanic (0 = No; 1= Yes);<br>NH White (0 = No; 1= Yes);<br>NH Black (0 = No; 1= Yes);<br>Other (0 = No; 1= Yes); |
| | R04R_A_HISP_IMP | PATH imputed Hispanic origin | 1 = Hispanic;<br>2 = Not Hispanic | |
| Education Attainment | R04R_A_AM0018_V2 | What is the highest grade or level of school you completed? (5 levels) | 1 = Less than High School<br>2 = GED<br>3 = High school graduate<br>4 = Some college (no degree) or associates degree<br>5 = Bachelor's degree or advanced degree | 1 = Less than high school<br>2 = GED<br>3 = High school graduate<br>4 = Some college (no degree) or associates degree<br>5 = Bachelor's degree or advanced degree |
| Annual Household Income (USD) | R04R_A_AM0030 | Which of the following categories best describes your total household income in the past 12 months? | 1 = Less than $10,000<br>2 = $10,000 to $24,999<br>3 = $25,000 to $49,999<br>4 = $50,000 to $99,999<br>5 = $100,000 or more | 0 = $50,000 or more;<br>1 = less than $10,000 - $49,999; |
| | R04_AM0031 | | 1 = Above $50,000<br>2 = Below $50,000 | |

| | | | | |
|---|---|---|---|---|
| | | Is your total household income during the past 12 months above or below $50,000? | | |
| Region | R04X_CB_REGION | Groupings of states and the District of Columbia that subdivide the United States for the presentation of census data. | 1 = Northeast<br>2 = Midwest<br>3 = South<br>4 = Wes | 1 = Northeast<br>2 = Midwest<br>3 = South<br>4 = West |
| Sexual Orientation | R04R_A_SEXORIENT2 | 2 category recode of sexual orientation. | 1 = Lesbian, Gay, Bisexual, Something else;<br>2 = Straight | 0 = Lesbian, Gay, Bisexual, Something else;<br>1 = Straight |
| Absolute Harm | R04_AC9050 | How harmful do you think cigarettes are to health? | 1 = Not at all harmful<br>2 = Slightly harmful<br>3 = Somewhat harmful<br>4 = Very harmful<br>5 = Extremely harmful | 1 = Not at all harmful<br>2 = Slightly harmful<br>3 = Somewhat harmful<br>4 = Very harmful<br>5 = Extremely harmful |
| Relative Harm | R04_AE1099 | Is using e-cigarettes or other electronic nicotine products less harmful, about the same, or more harmful than smoking cigarettes? | 1 = Less harmful<br>2 = About the same<br>3 = More harmful | 1 = Less harmful<br>2 = About the same<br>3 = More harmful |
| Household Policy of Cigarette Smoking | R04_AR1045 | For tobacco products that are burned, such as cigarettes, cigars, pipes or hookah, which statement best describes the rules about smoking a tobacco product inside your home? | 1 = It is not allowed anywhere or at any time inside my home<br>2 = It is allowed in some places or at some times inside my home<br>3 = It is allowed anywhere and at any time inside my home | 1 = It is not allowed anywhere or at any time inside my home<br>2 = It is allowed in some places or at some times inside my home<br>3 = It is allowed anywhere and at any time inside my home |
| Family Tobacco Use | R04R_A_AX0066 | Does anyone who lives with you now do any of the following: Smoke cigarettes, cigars, cigarillos or filtered cigars, pipe tobacco, use e-products exclusively, use other tobacco products, including smokeless, snus and hookah, or no one living in the home uses tobacco | 1 = Cigarettes, cigars, cigarillos or filtered cigars and pipe tobacco<br>2 = E-products exclusively<br>3 = Other tobacco products, including smokeless, snus and hookah<br>4 = No one living in the home uses tobacco | 0 = No one living in the home uses tobacco;<br>1= Cigarettes, cigars, cigarillos or filtered cigars/ Smokeless or other tobacco only |
| Internalization | R04_AX0161 | Last time you had significant problems with: Feeling very | 1 = Past month,<br>2 = 2 to 12 months ago, | = 0 if has less than 2 of (R04_AX0161, R04_AX0162, |

| | | | | |
|---|---|---|---|---|
| | | trapped, lonely, sad, blue, depressed or hopeless about the future | 3 = Over a year ago,<br>4 = Never | R04_AX0163, or R04_AX0164) in Past month or 2-12 months ago<br><br>=1 if had 2 or more of (R04_AX0161, R04_AX0162, R04_AX0163, or R04_AX0164) in Past month or 2-12 months ago; |
| | R04_AX0162 | Last time you had significant problems with: Sleep trouble - such as bad dreams, sleeping restlessly or falling asleep during the day | | |
| | R04_AX0163 | Last time you had significant problems with: Feeling very anxious, nervous, tense, scared, panicked or like something bad was going to happen | | |
| | R04_AX0164 | Last time you had significant problems with: Becoming very distressed and upset when something reminded you of the past | | |
| Externalization | R04_AX0165 | Last time you did the following two or more times: Lied or conned to get things you wanted or to avoid having to do something | 1 = Past month;<br>2 = 2 to 12 months ago;<br>3 = Over a year ago;<br>4 = Never | = 0 if has less than 2 of (R04_AX0165, R04_AX0166, R04_AX0167, R04_AX0168, R04_AX0169, R04_AX0250, or R04_AX0251) in Past month or 2-12 months ago<br><br>=1 if had 2 or more of (R04_AX0165, R04_AX0166, R04_AX0167, R04_AX0168, R04_AX0169, R04_AX0250, or R04_AX0251) in Past month or 2-12 months ago; |
| | R04_AX0166 | Last time you did the following two or more times: Had a hard time paying attention at school, work or home | | |
| | R04_AX0167 | Last time you did the following two or more times: Had a hard time listening to instructions at school, work or home | | |
| | R04_AX0168 | Last time you did the following two or more times: Were a bully or threatened other people | | |

| | | | | |
|---|---|---|---|---|
| | R04_AX0169, | Last time you did the following two or more times: Started physical fights with other people | | |
| | R04_AX0250 | Last time you did the following two or more times: Felt restless or the need to run around or climb on things | | |
| | R04_AX0251 | Last time you did the following two or more times: Gave answers before the other person finished asking the question | | |
| Heart Disease (P12M) | R04_AX0111_12M_01 | In past 12 months, doctor, nurse or other health professional said you had: High blood pressure | 1 = Marked<br>2 = Not Marked | 0 = No disease, indicate had 0 of (R04_AX0111_12M_01, R04_AX0111_12M_02, R04_AX0111_12M_03, R04_AX0111_12M_04, R04_AX0111_12M_05, or R04_AX0111_12M_06) in past 12 months<br><br>1 = have disease, indicate had at least 1 of (R04_AX0111_12M_01, R04_AX0111_12M_02, R04_AX0111_12M_03, R04_AX0111_12M_04, R04_AX0111_12M_05, or R04_AX0111_12M_06) in past 12 months<br><br>2 = missing values; indicate all missing of, R04_AX0111_12M_02, R04_AX0111_12M_03, R04_AX0111_12M_04, R04_AX0111_12M_05, or |
| | R04_AX0111_12M_02 | In past 12 months, doctor, nurse or other health professional said you had: High cholesterol | | |
| | R04_AX0111_12M_03 | In past 12 months, doctor, nurse or other health professional said you had: Congestive heart failure | | |
| | R04_AX0111_12M_04 | In past 12 months, doctor, nurse or other health professional said you had: A stroke | | |
| | R04_AX0111_12M_05 | In past 12 months, doctor, nurse or other health professional said you had: A heart attack (also called myocardial infarction) or needed bypass surgery | | |
| | R04_AX0111_12M_06 | In past 12 months, doctor, nurse or other health professional said you had: Some other heart condition | | |

| | | | | |
|---|---|---|---|---|
| | | | | R04_AX0111_12M_06) in past 12 months |
| Respiratory Disease (P12M) | R04_AX0119_12M_01 | In past 12 months, doctor, nurse or other health professional said you had a lung or respiratory condition: COPD | 1 = Marked<br>2 = Not Marked | 0 = No disease, indicate has 0 of (R04_AX0119_12M_01, R04_AX0119_12M_02, R04_AX0119_12M_03, R04_AX0119_12M_04, or R04_AX0119_12M_05) in the past 12 months |
| | R04_AX0119_12M_02 | In past 12 months, doctor, nurse or other health professional said you had a lung or respiratory condition: Chronic bronchitis | | |
| | R04_AX0119_12M_03 | In past 12 months, doctor, nurse or other health professional said you had a lung or respiratory condition: Emphysema | | 1 = have disease, indicate had at least 1 of (R04_AX0119_12M_01, R04_AX0119_12M_02, R04_AX0119_12M_03, R04_AX0119_12M_04, or R04_AX0119_12M_05) in past 12 months |
| | R04_AX0119_12M_04 | In past 12 months, doctor, nurse or other health professional said you had a lung or respiratory condition: Asthma | | 2 = missing values, indicate all missing of (R04_AX0119_12M_01, R04_AX0119_12M_02, R04_AX0119_12M_03, R04_AX0119_12M_04, or R04_AX0119_12M_05) in the past 12 months |
| | R04_AX0119_12M_05 | In past 12 months, doctor, nurse or other health professional said you had a lung or respiratory condition: Some other lung or respiratory condition | | |
| Cancer (P12M) | R04_AX0144_12M | In past 12 months, doctor, nurse or other health professional told you that you had cancer | 1 = Yes;<br>2 = No | 0 = No;<br>1 = Yes |
| BMI | R04R_A_BMI | Body mass index (BMI) | (Numerical) | 0 = [18.5, 25) Underweight<br>1 = (0, 18.5) Normal weight<br>2 = [25, 30) Overweight<br>3 = [30, ∞) Obesity |

| | | | | |
|---|---|---|---|---|
| Alcohol P30D Use | R04_AX0673 | Have you used alcohol within the past 30 days? | 1 = Yes; 2 = No | 0 = No; 1 = Yes |
| Marijuana P30D Use | R04_AX0675 | Have you used marijuana, hash, THC, grass, pot or weed within the past 30 days? | 1 = Yes; 2 = No | 0 = No; 1 = Yes |
| Other Combustible Tobacco Use | R04R_A_EVR_PIPE<br><br>R04R_A_EVR_HOOK<br><br>R04R_A_EVR_CIGAR | Wave 4 adult respondents who have ever smoked a pipe.<br><br>Wave 4 adult respondents who have ever smoked a hookah.<br><br>Wave 4 adult respondents who have ever smoked any cigar product. | 1 = Yes; 2 = No | 0 = No; 1 = Yes |
| Quit Intent | R04_AN0240 | Time-frame when planning to quit smoking/using tobacco product(s) for good | 1 = In the next 7 days<br>2 = In the next 30 days<br>3 = In the next 6 months<br>4 = In the next year<br>5 = More than one year from now | 0 = in the next 7 days - 6 months<br>1 = in the next year or more than one year from now |
| Secondhand Smoking | R04_AX0068 | During the past seven days, about how many hours were you around others who were smoking [whether or not you were smoking yourself]? Include time in your home, in a car, at work, or outdoors. | (Numerical) | (Numerical) |
| Time to First Smoking after Waking | R04_AC1024_NN<br><br>R04_AC1024_UN | Time to first cigarette after waking on days smoked - Number (current smokers)<br>Time to first cigarette after waking on days smoked - Unit (current smokers) | (Numerical)<br><br>1 = Minutes after waking<br>2 = Hours after waking | (Numerical) |
| Nicotine Dependence | R04_AN0025<br><br>R04_AN0030 | Level of agreement: Frequently crave tobacco product(s)<br><br>Level of agreement: Urges keep getting stronger if don't/Still have urges to smoke/use tobacco product(s) | 1 = Not true of me at all<br>2 = 2 on a scale of 1 to 5<br>3 = 3 on a scale of 1 to 5<br>4 = 4 on a scale of 1 to 5<br>5 = Extremely true of me | Rescale 1–5 to 0–100 scale:<br>1 → 0<br>2 or 3 →50<br>4 or 5 →100; |

| | | | |
|---|---|---|---|
| R04_AN0065 | Level of agreement: Can only go a couple of hours without smoking/using tobacco product(s) | | Then sum and count non-missing rescaled responses.<br><br>Finally Compute average per wave (scaled sum ÷ non-missing count) |
| R04_AN0035 | Level of agreement: Tobacco product(s) control me | | |
| R04_AN0045 | Level of agreement: My tobacco product(s) smoking/use is out of control/My urge to smoke/use tobacco product(s) is out of control | | |
| R04_AN0085 | Level of agreement: Would find it really hard to stop smoking/using tobacco product(s) | | |
| R04_AN0090 | Level of agreement: Would find it hard to stop smoking/using tobacco product(s) for a week | | |
| R04_AN0060 | Level of agreement: Usually want to smoke/use tobacco product(s) right after waking up | | |
| R04_AN0095 | Level of agreement: After not smoking/using tobacco product(s) for a while, need to smoke/use tobacco product(s) in order to feel less restless and irritable | | |
| R04_AN0100 | Level of agreement: After not smoking/using tobacco product(s) for a while, need to smoke/use tobacco product(s) in order to keep self from experiencing any discomfort | | |

| | | | | |
|---|---|---|---|---|
| | R04_AN0055 | Level of agreement: Finds self reaching for tobacco product(s) without thinking about it | | |
| | R04_AN0050 | Level of agreement: Frequently smoke/use tobacco product(s) without thinking about it | | |
| | R04_AN0070 | Level of agreement: Smoking/using tobacco product(s) really helps me feel better if feeling down | | |
| | R04_AN0075 | Level of agreement: Smoking/using tobacco product(s) helps me think better | | |
| | R04_AN0080 | Level of agreement: Would feel alone without my tobacco product(s) | | |
| E-cigarette Use | R04R_A_P30D_EPRODS | Wave 4 adult past-30-day electronic nicotine product use | 1 = Yes; 2 = No | 0 = No use<br>1 = Some-day use<br>2 = Daily use |
| | R04_AV10003 | Do you now use electronic nicotine products | 1 = Every day<br>2 = Some days<br>3 = Not at all | |
| Cigarette Smoking Days (Frequency) | R04R_A_P30D_CIGS | Have smoked in the past 30 days. | 1 = Yes; 2 = No | 0 - 30 |
| | R04_AC1003 | Do you now smoke cigarettes? | 1 = Every day<br>2 = Some days<br>3 = Not at all | |
| | R04_AC1022 | How many of the past 30 days smoked a cigarette? | (numeric) | |
| | | **Exposure (Wave 5)** | | |
| E-cigarette Use | R05R_A_P30D_EPRODS | Wave 5 adult past-30-day electronic nicotine product use | 1 = Yes; 2 = No | 0 = No use<br>1 = Some-day use<br>2 = Daily use |

| | | | | |
|---|---|---|---|---|
| (Days of e-cigarette use in the past 30 days) | R05_AV1003 | Do you now use electronic nicotine products | 1 = Every day<br>2 = Some days<br>3 = Not at all | |
| | | **Outcome (Wave 6)** | | |
| Cigarette Smoking Days (Frequency) | R06R_A_P30D_CIGS | Have smoked in the past 30 days. | 1 = Yes; 2 = No | 0 - 30 |
| | R06_AC1003 | Do you now smoke cigarettes? | 1 = Every day<br>2 = Some days<br>3 = Not at all | |
| | R06_AC1022 | How many of the past 30 days smoked a cigarette? | (numeric) | |

Table S2. Characteristics of the study sample at Wave 4 and by Exposure Groups

| Variable | Whole Sample (n = 4909, %) | Weighted (%) | E-cigarette No Use (n = 4032, 82.1%) | E-cigarette Some-Day Use (n = 606, 12.3%) | E-cigarette Daily Use (n = 271, 5.5%) |
|---|---|---|---|---|---|
| **Age Group** | | | | | |
| 18 to 24 years | 643 (13.1%) | 8.4 | 410 (10.1%) | 144 (23.8%) | 89 (32.8%) |
| 25 to 34 years | 1170 (23.8%) | 23.8 | 902 (22.4%) | 184 (30.4%) | 84 (31.0%) |
| 35 to 44 years | 902 (18.4%) | 20.3 | 744 (18.5%) | 113 (18.6%) | 45 (16.6%) |
| 45 to 54 years | 1009 (20.6%) | 21.3 | 873 (21.7%) | 106 (17.5%) | 30 (11.1%) |
| 55 to 64 years | 869 (17.7%) | 18.8 | 812 (20.1%) | 40 (6.6%) | 17 (6.3%) |
| 65+ years | 316 (6.4%) | 7.4 | 291 (7.2%) | 19 (3.1%) | 6 (2.2%) |
| **Sex** | | | | | |
| Female | 2667 (54.2%) | 47.3 | 2195 (54.4%) | 340 (56.1%) | 132 (48.7%) |
| Male | 2242 (45.8%) | 52.7 | 1837 (45.6%) | 266 (43.9%) | 139 (51.3%) |
| **Race/Ethnicity** | | | | | |
| Hispanic | 639 (13.0%) | 12.4 | 530 (13.2%) | 87 (14.4%) | 22 (8.1%) |
| NH White | 3081 (62.8%) | 67.7 | 2489 (61.7%) | 384 (63.4%) | 208 (76.8%) |
| NH Black | 850 (17.3%) | 14.5 | 748 (18.6%) | 82 (13.5%) | 20 (7.4%) |
| Others | 338 (6.9%) | 5.5 | 264 (6.5%) | 53 (8.7%) | 21 (7.7%) |
| Missing | 1 (0.0%) | 0.0 | 1 (0.0%) | 0 (0.0%) | 0 (0.0%) |
| **Education Attainment** | | | | | |
| Less than high school | 853 (17.4%) | 16.0 | 716 (17.8%) | 94 (15.5%) | 43 (15.9%) |
| GED | 598 (12.2%) | 11.7 | 507 (12.6%) | 68 (11.2%) | 23 (8.5%) |
| High school graduate | 1269 (25.9%) | 31.0 | 1053 (26.1%) | 147 (24.3%) | 69 (25.5%) |
| Some college | 1676 (34.1%) | 31.5 | 1337 (33.2%) | 228 (37.6%) | 111 (41.0%) |
| Bachelor's or higher degree | 495 (10.1%) | 9.4 | 402 (10%) | 69 (11.4%) | 24 (8.9%) |
| Missing | 18 (0.3%) | 0.4 | 17 (0.3%) | 0 (0.0%) | 1 (0.4%) |
| **Annual Household Income** | | | | | |
| 0 - $50,000 | 1140 (23.2%) | 26.4 | 922 (22.9%) | 141 (23.3%) | 77 (28.4%) |
| >= $50,000 | 3640 (74.2%) | 70.8 | 3003 (74.5%) | 451 (74.4%) | 186 (68.6%) |
| Missing | 129 (2.6%) | 2.8 | 107 (2.6%) | 14 (2.3%) | 8 (3.0%) |
| **Region** | | | | | |
| Northeast | 704 (14.3%) | 16.6 | 580 (14.4%) | 83 (13.7%) | 41 (15.2%) |
| Midwest | 1391 (28.3%) | 24.2 | 1133 (28.1%) | 183 (30.2%) | 75 (27.7%) |
| South | 1959 (39.9%) | 41.8 | 1618 (40.1%) | 231 (38.1%) | 110 (40.6%) |
| West | 855 (17.5%) | 17.4 | 701 (17.4%) | 109 (18.0%) | 45 (16.6%) |
| **Sexual Orientation** | | | | | |
| Not straight | 496 (10.1%) | 8.6 | 346 (8.6%) | 102 (16.8%) | 48 (17.7%) |
| Straight | 4381 (89.2%) | 90.7 | 3658 (90.7%) | 503 (83.0%) | 220 (81.2%) |
| Missing | 32 (0.7%) | 0.7 | 28 (0.7%) | 1 (0.2%) | 3 (1.1%) |
| **Absolute Harm Perception** | | | | | |
| Not at all harmful | 100 (2.0%) | 2 | 72 (1.8%) | 19 (3.1%) | 9 (3.2%) |
| Slightly harmful | 248 (5.1%) | 4.9 | 203 (5.0%) | 25 (4.1%) | 20 (7.4%) |
| Somewhat harmful | 1119 (22.8%) | 23 | 927 (23.0%) | 145 (23.9%) | 47 (17.3%) |
| Very harmful | 1779 (36.2%) | 37 | 1453 (36.0%) | 227 (37.5%) | 99 (36.5%) |
| Extremely harmful | 1657 (33.8%) | 32.9 | 1373 (34.1%) | 190 (31.4%) | 94 (34.7%) |
| Missing | 6 (0.1%) | 0.2 | 4 (0.1%) | 0 (0.0%) | 2 (0.7%) |
| **Relative Harm Perception** | | | | | |
| Less harmful | 1065 (21.7%) | 21.3 | 770 (19.1%) | 176 (29.0%) | 119 (43.9%) |
| About the same | 3323 (67.7%) | 68.3 | 2809 (69.7%) | 380 (62.7%) | 134 (49.4%) |

| Variable | Whole Sample (n = 4909, %) | Weighted (%) | E-cigarette No Use (n = 4032, 82.1%) | E-cigarette Some-Day Use (n = 606, 12.3%) | E-cigarette Daily Use (n = 271, 5.5%) |
|---|---|---|---|---|---|
| More harmful | 434 (8.8%) | 8.6 | 377 (9.4%) | 43 (7.1%) | 14 (5.2%) |
| Missing | 87 (1.8%) | 1.9 | 76 (1.8%) | 7 (1.2%) | 4 (1.5%) |
| **House Policy of Cigarette Smoking** | | | | | |
| not allowed anywhere/anytime | 2562 (52.1%) | 53.7 | 2074 (51.4%) | 318 (52.5%) | 170 (62.7%) |
| allowed in some places or at sometimes | 1230 (25.1%) | 23.8 | 1021 (25.3%) | 158 (26.1%) | 51 (18.8%) |
| allowed in anywhere/at any time | 1099 (22.4%) | 22.2 | 919 (22.8%) | 130 (21.5%) | 50 (18.5%) |
| Missing | 18 (0.4%) | 0.3 | 18 (0.4%) | 0 (0.0%) | 0 (0.0%) |
| **Family Tobacco Use** | | | | | |
| No | 1507 (30.7%) | 31.5 | 1245 (30.9%) | 181 (29.9%) | 81 (29.9%) |
| Yes | 2551 (52.0%) | 49.7 | 2048 (50.8%) | 351 (57.9%) | 152 (56.1%) |
| Missing | 851 (17.3%) | 18.8 | 739 (18.3%) | 74 (12.2%) | 38 (14.0%) |
| **Internalization** | | | | | |
| No | 2394 (48.8%) | 51.3 | 2050 (50.8%) | 249 (41.1%) | 95 (35.1%) |
| Yes | 2505 (51.0%) | 48.4 | 1973 (48.9%) | 356 (58.7%) | 176 (64.9%) |
| Missing | 10 (0.2%) | 0.2 | 9 (0.3%) | 1 (0.2%) | 0 (0.0%) |
| **Externalization** | | | | | |
| No | 2972 (60.5%) | 62.2 | 2557 (63.4%) | 300 (49.5%) | 115 (42.4%) |
| Yes | 1927 (39.3%) | 37.5 | 1465 (36.3%) | 306 (50.5%) | 156 (57.6%) |
| Missing | 10 (0.2%) | 0.3 | 10 (0.3%) | 0 (0.0%) | 0 (0.0%) |
| **Heart Disease in P12M[a]** | | | | | |
| No | 1673 (34.1%) | 33.8 | 1346 (33.4%) | 231 (38.1%) | 96 (35.4%) |
| Yes | 1129 (23.0%) | 23.9 | 984 (24.4%) | 103 (17.0%) | 42 (15.5%) |
| Missing | 2107 (42.9%) | 42.3 | 1702 (42.2%) | 272 (44.9%) | 133 (49.1%) |
| **Respiratory Disease in P12M[a]** | | | | | |
| No | 2176 (44.3%) | 45.5 | 1819 (45.2%) | 253 (41.7%) | 104 (38.4%) |
| Yes | 625 (12.8%) | 12.1 | 510 (12.6%) | 81 (13.4%) | 34 (12.5%) |
| Missing | 2108 (42.9%) | 42.4 | 1703 (42.2%) | 272 (44.9%) | 133 (49.1%) |
| **Cancer in P12M[a]** | | | | | |
| No | 2737 (55.8%) | 56.1 | 2277 (56.5%) | 326 (53.8%) | 134 (49.4%) |
| Yes | 66 (1.3%) | 1.5 | 56 (1.4%) | 7 (1.2%) | 3 (1.1%) |
| Missing | 2106 (42.9%) | 42.4 | 1699 (42.1%) | 273 (45.0%) | 134 (49.4%) |
| **BMI** | | | | | |
| Underweight | 115 (2.3%) | 2.3 | 98 (2.4%) | 11 (1.8%) | 6 (2.2%) |
| Normal Weight | 1561 (31.8%) | 32.2 | 1249 (31.0%) | 215 (35.5%) | 97 (35.8%) |
| Overweight | 1508 (30.7%) | 32.3 | 1264 (31.3%) | 172 (28.4%) | 72 (26.6%) |
| Obesity | 1643 (33.5%) | 31.5 | 1357 (33.7%) | 199 (32.8%) | 87 (32.1%) |
| Missing | 82 (1.7%) | 1.8 | 64 (1.6%) | 9 (1.5%) | 9 (3.3%) |
| **Alcohol Use in P30D[b]** | | | | | |
| No | 2018 (41.1%) | 40.0 | 1683 (41.7%) | 223 (36.8%) | 112 (41.3%) |
| Yes | 2801 (57.1%) | 58.2 | 2278 (56.5%) | 370 (61.1%) | 153 (56.5%) |
| Missing | 90 (1.8%) | 1.8 | 71 (1.8%) | 13 (2.1%) | 6 (2.2%) |
| **Marijuana Use in P30D[b]** | | | | | |
| No | 3218 (65.6%) | 67.8 | 2742 (68.0%) | 316 (52.1%) | 160 (59.0%) |
| Yes | 1479 (30.1%) | 28.2 | 1107 (27.5%) | 268 (44.2%) | 104 (38.4%) |
| Missing | 212 (4.3%) | 4.0 | 183 (4.5%) | 22 (3.6%) | 7 (2.6%) |

| Variable | Whole Sample (n = 4909, %) | Weighted (%) | E-cigarette No Use (n = 4032, 82.1%) | E-cigarette Some-Day Use (n = 606, 12.3%) | E-cigarette Daily Use (n = 271, 5.5%) |
|---|---|---|---|---|---|
| **Other Combustible Tobacco Use** | | | | | |
| No | 1083 (22.0%) | 22.1 | 991 (24.5%) | 61 (10.1%) | 31 (11.4%) |
| Yes | 2693 (54.9%) | 56.7 | 2232 (55.4%) | 319 (52.6%) | 142 (52.4%) |
| Missing | 1133 (23.1%) | 21.2 | 809 (20.1%) | 226 (37.3%) | 98 (36.2%) |
| **Quit Intent** | | | | | |
| No | 2513 (51.2%) | 50.3 | 2062 (51.2%) | 316 (52.1%) | 135 (49.8%) |
| Yes | 1265 (25.8%) | 25.7 | 1038 (25.7%) | 144 (23.8%) | 83 (30.6%) |
| Missing | 1131 (23.0%) | 24.0 | 932 (23.1%) | 146 (24.1%) | 53 (19.6%) |
| **Second-hand Smoking Exposure in P7D[c] (in hours)** | | | | | |
| Mean (SD) | 16.84 (27.67) | 16.33 (0.46[d]) | 16.74 (27.80) | 17.88 (28.46) | 16.04 (23.77) |
| **Time to First Smoking after Waking** | | | | | |
| Mean (SD) | 79.81 (150.15) | 80.06 (2.52[d]) | 77.74 (148.94) | 92.26 (159.30) | 82.77 (146.00) |
| **Nicotine dependence** | | | | | |
| Mean (SD) | 55.98 (28.16) | 55.92 (0.44[d]) | 56.39 (28.04) | 54.67 (28.00) | 52.77 (30.12) |
| **E-cigarette Use** | | | | | |
| No Use | 4208 (85.7%) | 86.7 | 3663 (90.7%) | 408 (67.3%) | 137 (50.6%) |
| Some-day use | 523 (10.7%) | 9.5 | 308 (7.6%) | 154 (25.4%) | 61 (22.5%) |
| Daily Use | 168 (3.4%) | 3.6 | 52 (1.3%) | 43 (7.1%) | 73 (26.9%) |
| Missing | 10 (0.2%) | 0.2 | 9 (0.2%) | 1 (0.2%) | 0.0 (0.0%) |
| **Smoking Day** | | | | | |
| Mean (SD) | 26.89 (7.64) | 26.85 (0.135[d]) | 27.18 (7.35) | 25.89 (8.66) | 24.86 (8.86) |

Notes. ENDS Exposure were measured at Wave 5.

[a] P12M = past 12 months, [b] P30D = past 30 days, [c] P7D = past 7 days, [d] standard errors.

Table S3 Causal Effects ENDS Use on Subsequent Smoking Days: Estimates of ATE and Group-Specific ATE by Group (weighted)

| | Daily Use vs. No Use | | | Some-Day Use vs. No Use | | | Daily Use vs. Some-Day Use | | |
|---|---|---|---|---|---|---|---|---|---|
| | estimate | s.e. | 95% CI | estimate | s.e. | 95% CI | estimate | s.e. | 95% CI |
| **ATE (Overall)** | -6.15 | 1.36 | (-8.10, -5.29) | -0.96 | 0.48 | (-1.56, -0.31) | -6.54 | 0.88 | (-7.35, -5.04) |
| **Group-Specific ATE** | | | | | | | | | |
| **Sex** | | | | | | | | | |
| Male | -6.24 | 1.43 | (-8.34, -5.44) | -0.85 | 0.49 | (-1.47, -0.19) | -6.75 | 0.88 | (-7.57, -5.33) |
| Female | -6.08 | 1.33 | (-7.93, -5.14) | -1.06 | 0.50 | (-1.64, -0.38) | -6.35 | 0.90 | (-7.15, -4.79) |
| **Race/Ethnicity** | | | | | | | | | |
| NH White | -6.33 | 1.32 | (-8.17, -5.44) | -0.87 | 0.51 | (-1.57, -0.15) | -6.55 | 0.85 | (-7.34, -5.15) |
| Hispanic | -5.46 | 1.49 | (-7.64, -4.65) | -1.31 | 0.64 | (-2.07, -0.47) | -5.20 | 1.09 | (-6.48, -3.72) |
| NH Black | -5.78 | 1.56 | (-7.97, -4.81) | -0.95 | 0.55 | (-1.58, -0.29) | -6.08 | 1.18 | (-7.25, -4.02) |
| Other | -6.87 | 1.54 | (-8.96, -6.04) | -1.08 | 0.57 | (-1.83, -0.29) | -8.99 | 0.91 | (-9.69, -7.56) |
| **Annual Household Income** | | | | | | | | | |
| <$50,000 | -6.13 | 1.37 | (-8.10, -5.23) | -0.76 | 0.47 | (-1.34, -0.13) | -6.69 | 0.91 | (-7.54, -5.15) |
| >=$50,000 | -6.22 | 1.38 | (-8.14, -5.36) | -1.49 | 0.56 | (-2.27, -0.76) | -6.73 | 0.83 | (-7.48, -5.33) |
| **Internalization** | | | | | | | | | |
| Yes | -6.19 | 1.38 | (-8.10, -5.40) | -0.88 | 0.48 | (-1.45, -0.25) | -6.09 | 0.87 | (-7.08, -4.67) |
| No | -6.11 | 1.37 | (-8.09, -5.17) | -1.05 | 0.52 | (-1.77, -0.33) | -7.23 | 0.94 | (-7.94, -5.62) |
| **Externalization** | | | | | | | | | |
| Yes | -6.27 | 1.28 | (-8.02, -5.59) | -1.46 | 0.52 | (-2.13, -0.80) | -6.08 | 0.85 | (-6.99, -4.71) |
| No | -6.08 | 1.43 | (-8.14, -5.09) | -0.65 | 0.49 | (-1.29, -0.02) | -7.04 | 0.94 | (-7.87, -5.47) |
| **Sexual Orientation** | | | | | | | | | |
| Straight | -6.19 | 1.37 | (-8.14, -5.31) | -0.98 | 0.49 | (-1.60, -0.31) | -6.48 | 0.88 | (-7.23, -4.98) |
| Not Straight | -5.79 | 1.40 | (-7.65, -5.06) | -0.78 | 0.51 | (-1.41, -0.19) | -6.82 | 0.93 | (-7.74, -5.34) |
| **Age Group (in years)** | | | | | | | | | |
| 18 - 24 | -6.28 | 1.37 | (-8.03, -5.68) | -0.24 | 0.44 | (-0.88, 0.28) | -6.32 | 0.84 | (-7.18, -5.02) |
| 25 - 34 | -6.66 | 1.42 | (-8.71, -5.91) | -0.96 | 0.53 | (-1.66, -0.36) | -6.24 | 0.89 | (-7.13, -4.81) |
| 35 - 44 | -5.99 | 1.30 | (-7.90, -5.34) | -1.09 | 0.52 | (-1.64, -0.32) | -7.32 | 1.02 | (-8.17, -5.68) |
| 45 - 54 | -5.50 | 1.45 | (-7.52, -4.33) | -0.70 | 0.54 | (-1.37, 0.09) | -6.44 | 1.11 | (-7.32, -4.55) |
| 55 - 64 | -6.37 | 1.45 | (-8.43, -5.12) | -1.39 | 0.64 | (-2.14, -0.50) | -7.74 | 1.06 | (-8.62, -6.10) |
| >= 65 | -6.06 | 1.53 | (-8.50, -5.02) | -1.57 | 0.85 | (-2.68, -0.33) | -4.56 | 1.12 | (-5.90, -2.88) |

Notes. ATE = average treatment/exposure effect; NH White = Non-Hispanic White; NH Black = Non=Hispanic Black; s.e. = standard error; CI = credible intervals.

Table S4 Residual Distribution Statistics BART for ATE effects

| | n | M | SD | Skewness |
|---|---|---|---|---|
| Daily vs No Use | 4301 | 0.01 | 10.04 | -1.28 |
| Some-Day vs No Use | 877 | 0.36 | 11.21 | -0.58 |
| Daily vs Some-Day Use | 4636 | -0.01 | 9.94 | -1.37 |

Figure S1 Flowchart of sample selection

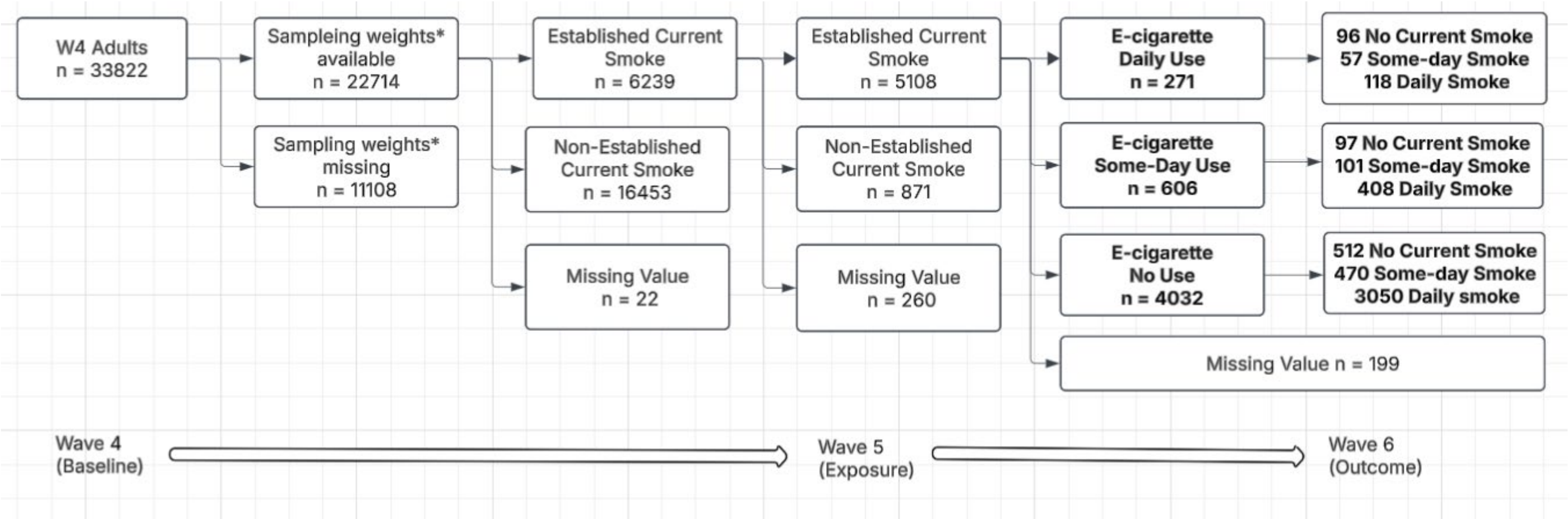


* Longitudinal Wave 4–6 sampling weights (W4 cohort longitudinal sampling weights at Wave 6; R06_A_S04WGT), a PATH Study derived sampling variable, were applied to account for the PATH Study's complex survey design, unequal probabilities of selection, differential nonresponse, and attrition between waves. Variance estimates were computed using the PATH balanced repeated replication (BRR) replicate weights (R06_A_S04WGT1 - R06_A_S04WGT100) with Fay's adjustment.

Boldface indicates the study sample.

Figure S2 Contour Plots of Sensitivity to unmeasured confounding for ATEs

| Exposure Contrast | Contour Plot |
|---|---|
| Daily vs No Use | 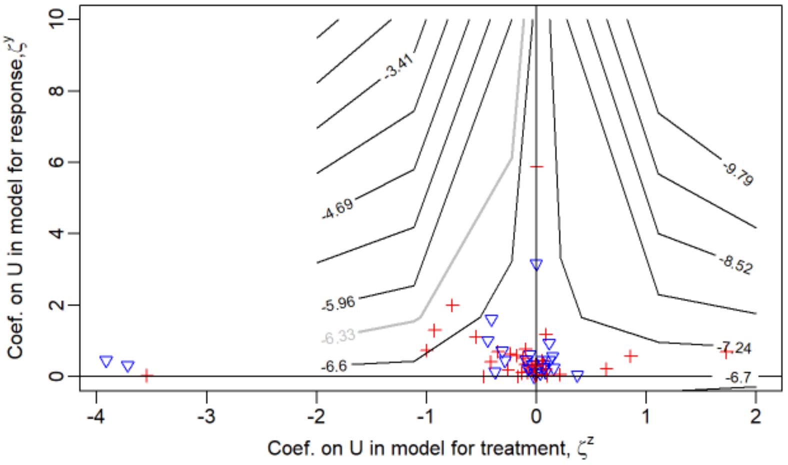 |
| Some-Day vs No | 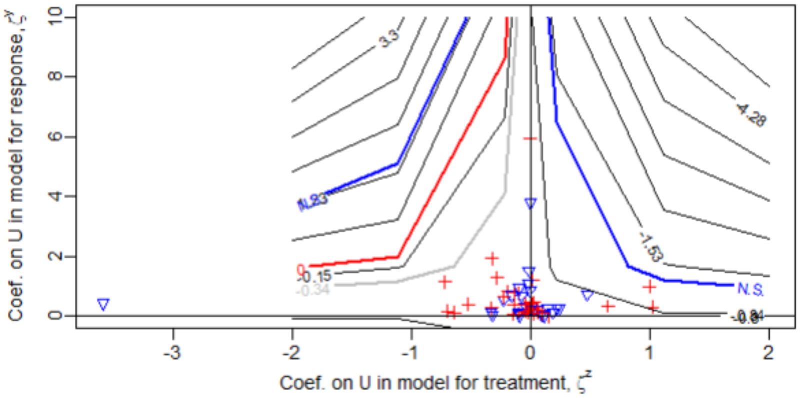 |
| Daily vs Some-Day | 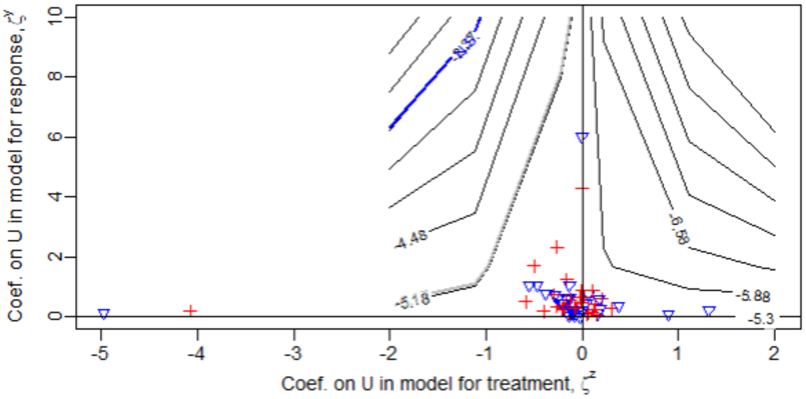 |

Each panel illustrates the sensitivity of the estimated treatment effect to unmeasured confounding for one exposure comparison: daily vs. no ENDS use, some-day vs. no ENDS use, and daily vs. some-day ENDS use. The analysis addresses the question: How strong would an unmeasured confounder need to be to explain away the estimated exposure effect? In each contour plot, the x-axis represents the association between a hypothetical unmeasured confounder and the ENDS exposure assignment, whereas the y-axis represents its association with the smoking days outcome. Larger absolute values indicate a stronger unmeasured confounder. Each contour represents the estimated treatment effect after adjusting for a hypothetical confounder with the corresponding exposure and outcome associations. The red "+" symbols and blue triangles represent the observed covariates, with their coordinates indicating the strength of each covariate's association with treatment assignment (x-axis) and smoking days (y-axis). The gray curve denotes the zero-bias boundary, where a hypothetical confounder would be sufficiently strong to explain away the observed treatment effect. Observed covariates located farther from this boundary indicate greater robustness of the estimated causal effect to unmeasured confounding.

**Appendix**. An introduction to Bayesian Additive Regression Tree for Causal Inference: a Causal Machine Learning Method

*Machine learning*, including tree-based models, is a broad field focused on developing algorithms that learn patterns from data to make predictions. A decision tree is a single predictive model that recursively partitions the data into increasingly homogeneous groups based on predictor variables. Decision trees are easy to interpret; however, they are often unstable because small changes in the data can result in substantially different tree structures. Bagging (or Bootstrap Aggregating) improves prediction by fitting many decision trees to different bootstrap samples of the data and averaging their predictions, thereby reducing variance and improving stability. Boosting also combines multiple trees, but instead of constructing them independently, it builds them sequentially, with each new tree focusing on the prediction residuals from the previous trees. As a result, the final prediction is the sum of a series of small trees, with each tree making a modest contribution to the overall model. This sequential approach primarily reduces bias and often achieves higher predictive accuracy than decision tree or bagging. Bayesian Additive Regression Trees (BART) is a Bayesian ensemble tree method. Like boosting, BART represents the prediction as the sum of many small decision trees, each tree explains a modest proportion of the residual variation left unexplained by the previous tree(s).

*BART*. BART is an ensemble method that represent an unknown function $f(x)$ as the sum of $m$ distinct regression trees. The "sum-of-tree" model for a response Y is expressed as:

$$Y = \sum_{j=1}^{K} g(x; T_j, M_j) + \varepsilon,\ \varepsilon \sim N(0, \sigma^2).$$

In this expression, each $g(x; T_j, M_j)$ represent a single regression tree $j$ with its structure $T_j$ and leaf parameters $M_j$. The final prediction is the cumulative contribution of all m trees plus residual. The core of the BART algorithm is the Markov chain Monte Carlo (MCMC) procedure, in which the ensemble of trees is updated iteratively to approximate the posterior distribution of the unknown regression function. BART defines prior distributions: prior on tree structure ($P(split\ at\ depth\ d) = 0.95(1+d)^{-2}$), prior on terminal node means (each leaf parameter has $\mu_k \sim N(0, (\frac{0.5}{K\sqrt{m}})^2)$, and prior on residual variance ($\sigma^2 \sim Inverse\ \chi^2$). Hence the posterior distribution is $P(T, M, \sigma^2) \propto p(T, M, \sigma^2)P(T)P(M)P(\sigma^2)$. Rather than constructing trees sequentially as in boosting, through many MCMC iterations, BART updates one tree at a time while conditioning on the current values of all the remaining trees.

*MCMC iteration*. At the beginning of each MCMC iteration, all K trees have an initial structure and a set of terminal node predictions. To update the j$^{th}$ tree, the algorithm temporarily holds the other K−1 trees fixed and computes the partial residuals, $\varepsilon = y_i - \sum_{j=1}^{K} g(x; T_j, M_j)$. Updating a tree involves two steps. First, a new tree structure is proposed by making a small modification to the current tree, such as growing a terminal node, pruning a branch, changing a splitting rule, or swapping parent and child splitting rules. The proposed tree is accepted or rejected according to a Metropolis-Hastings criterion, which favors models with higher posterior probability while respecting the regularization priors that encourage shallow trees. Once the tree structure has been determined, the terminal node means are sampled from their full conditional posterior distributions. After all K trees have been updated in this manner, the residual variance $\sigma^2$ is sampled from its posterior distribution. This completes one MCMC iteration.

In our analyses, the algorithm repeats this process for 700 of iterations. Early 200 iterations (the burn-in period) are discarded to allow the Markov chain to converge to the target posterior distribution. Each subsequent iteration (500 burn-out) produces one plausible ensemble of regression trees. Rather than selecting a single "best" model, BART averages predictions across all retained posterior samples, enabling both accurate prediction and direct quantification of uncertainty through posterior means (e.g., ATE) and credible intervals (e.g., 95% CI for the ATE). In this study, we repeated the estimation procedure 100 times using the PATH replicate weights and constructed 95% CI from the empirical distribution of the resulting estimates, following guidance from the PATH Technical Working Group.

*Causal Machine Learning Methods*. Causal machine learning methods implement flexible machine learning algorithms, such as Bayesian Additive Regression Trees (BART), random forest, gradient boosting, or Super Learners, which can automatically capture nonlinearities and high-order interactions while reducing the need for manual model specification. Despite these advantages over conventional perimetric models, causal machine learning methods do not relax the fundamental assumptions required for causal inference. Both traditional and machine learning–based causal inference approaches require no unmeasured confounding, positivity (nonzero probability of receiving each treatment), Stable Unit Treatment Value Assumption (consisting of consistency and no interference), and correct temporal ordering. In other words, machine learning cannot recover causal effects if important confounders are omitted or if the causal assumptions are violated. Rather, its advantage lies in reducing bias due to model misspecification and improving estimation of nuisance functions, which can lead to more accurate treatment effect estimates, especially in high-dimensional settings.

Many modern causal inference methods, such as doubly robust estimation, targeted maximum likelihood estimation (TMLE), double/debiased machine learning (DML), and causal forests, combine machine learning with semiparametric theory to achieve valid statistical inference while retaining the flexibility of machine learning. Consequently, causal machine learning should be viewed not as a replacement for traditional causal inference, but as an extension that enhances the estimation of causal effects when the underlying relationships are complex.
